# Cost-effectiveness of respiratory syncytial virus vaccination strategies for older Canadian adults: a multi-model comparison

**DOI:** 10.1101/2024.05.16.24307501

**Authors:** Monica Rudd, Alison E. Simmons, Gebremedhin B. Gebretekle, Ashleigh R. Tuite

## Abstract

**Background:** Two respiratory syncytial virus (RSV) vaccines are currently approved for use in adults aged 60 years and older in Canada. Economic analyses have shown that adult RSV vaccination programs may be cost-effective, particularly when focused on people at increased risk of RSV disease due to increased age or presence of chronic medical conditions (CMCs). We conducted a multi-model comparison to explore the impact of alternate model structural and methodological assumptions on the cost-effectiveness of RSV vaccination programs.

**Methods:** We compared three static cost-utility models developed by the Public Health Agency of Canada, GSK, and Pfizer using a common set of input parameters. Each model evaluated sequential incremental cost-effectiveness ratios (ICERs) in 2023 Canadian dollars per quality-adjusted life year (QALY) for a set of policy alternatives, with vaccine eligibility determined by combinations of age and CMC status. Results were calculated for each vaccine separately for scenarios assuming two or three years of vaccine protection using the health system perspective and a 1.5% annual discount rate.

**Results:** The three cost-utility models were broadly concordant across the scenarios modeled. In all scenarios, focusing on vaccination of people with CMCs was preferred over broader age-based policies. RSV vaccination for people with CMCs over the age of 70 years was most commonly identified as the optimal policy when using a cost-effectiveness threshold of $50,000/QALY. When only considering policies based on age criteria, vaccinating people over 80 years was cost-effective at this threshold.

**Discussion:** A multi-model comparison of Canadian cost-utility models shows that RSV vaccination programs for RSV are likely cost-effective for some groups of older adults in Canada. These findings were consistent across models, despite differences in model structure.

## BACKGROUND

Respiratory syncytial virus (RSV) is a major cause of respiratory infections in Canada, with a large burden of disease occurring in young children and older adults (1). RSV was estimated to account for 4.8% of hospitalizations for acute respiratory infections among Canadian adults aged over 50 years between 2012 and 2015 (2). Hospital mortality rates increased with age and among those with chronic medical conditions (2-4).

With the recent authorization of two vaccines for adults aged over 60 years in Canada, policy-makers are evaluating the use of these products in this population, including whether to recommend publicly-funded vaccination programs (5). Economic considerations are one important input to these decision processes.

We recently conducted an analysis of the cost-effectiveness of various vaccination program options for older adults in Canada (6) to inform forthcoming National Advisory Committee on Immunization (NACI) recommendations for the use of RSV vaccines in older adults. This analysis showed that vaccinating older adults may be cost-effective, depending on the program design. In particular, we showed that programs focused on vaccinating people with chronic medical conditions (CMCs) that place them at increased risk of RSV disease are expected to provide better value for money than more general age-based programs.

While model-based economic evaluations can provide useful insights for decision-makers, an exploration of how uncertainty impacts the results is important to avoid making suboptimal decisions. Sensitivity analyses can be performed to test uncertainty due to model inputs and parameter assumptions. Although changes to assumptions about model structure can be assessed in scenario analyses, such analyses may be challenging. Multi-model comparison studies can be used to address uncertainty due to model structure and methodology and are recommended in the NACI guidelines for economic evaluations (7-9). By comparing results across independently-developed economic models with standardized input parameters, researchers can assess the extent to which model-derived results are robust to differences in model mathematical formulation and methodological choices, allowing for higher confidence when evaluating this evidence.

We conducted a multi-model comparison of three economic cost-utility models to assess the robustness of findings about the cost-effectiveness of RSV vaccination program options in older adults in Canada to variation in model assumptions and structure.

## METHODS

### Model Selection

The multi-model comparison was conducted to support the National Advisory Committee on Immunization (NACI) and was part of an economic evidence package that considered during the development of recommendations for the use of RSV vaccines in Canadian adults. In addition to the Public Health Agency of Canada (PHAC)-developed model, we restricted our focus to models from manufacturers with a product approved for use in Canadian adults aged over 60 years for the 2024-2025 RSV season (Arexvy (GSK) and Abrysvo (Pfizer)). GSK and Pfizer provided their models, which were both constructed in Microsoft Excel (10). Model reparameterization and re-analyses were conducted by our team.

### Health Economic Framework

We evaluated all models in a population of 100,000 Canadian adults over the age of 50 years. Although the current RSV vaccines were authorized for use in the population age 60 years and older at the time of the analysis, we included some vaccination strategies that considered a lower age limit of 50 years, given that a lower age indication is currently under review (11). The population was distributed by age group (12) and stratified as higher risk or average risk based on the presence or absence of any chronic medical conditions (CMCs) placing them at increased risk of RSV disease (13). Model-estimated outcomes of interest included the number of RSV-attributable outpatient healthcare provider visits, emergency department visits, hospitalizations, deaths, and adverse events following immunization, QALY losses, vaccination costs, and healthcare costs. We computed expected vaccination costs, RSV-attributable medical costs, and quality-adjusted life year (QALY) losses for a range of possible vaccination programs. We evaluated the impact of vaccination programs using either Arexvy or Abrysvo, for a policy time horizon of two to three years, depending on the assumed duration of vaccine protection. All models began in September of the first year to cover the expected start of the typical RSV season (prior to the SARS-CoV-2 pandemic), with vaccination occurring at the start of the first season. Lifetime QALY losses were computed in the case of RSV mortality. All costs and QALY losses were discounted at a rate of 1.5% per annum (9). Costs and QALYs were used to compute sequential incremental cost-effectiveness ratios (ICERs) for all policy options under consideration. We only considered the health system perspective in this analysis.

### Model Overviews and Standardization

Each cost-utility model had unique features that required us to adapt input assumptions to be directly comparable, as described below. An overview of important characteristics of the PHAC, GSK, and Pfizer models is also provided in **Table 1**.

**Table 1.** Overview of models included in the multi-model comparison.

| Model Attribute | PHAC | GSK | Pfizer |
| --- | --- | --- | --- |
| Dynamics | Static | Static | Static |
| Aggregation | Individual | Cohort | Cohort |
| Age groups (years) | 50-59, 60-64, 65-69, 70-74, 75-79, 80+ | 50-59, 60-64, 65-69, 70-74, 75-79, 80+ | 18-49<sup>a</sup>, 50-59, 60-69, 70-79, 80+ |
| Risk strata | Two strata representing people with and without chronic medical conditions | Strata not explicitly modeled. Average and high risk adults were modeled as separate cohorts | Two or three risk strata, with option to allow people to move to higher risk strata as they age |
| RSV-related outcomes | Outpatient healthcare provider visit, ED visit, hospitalization, ICU | RSV-URTD<sup>b</sup> (outpatient healthcare provider or ED visit), RSV-LRTD<sup>c</sup> (hospitalization) | Non-medically attended, outpatient visit, ED visit, hospitalization |
| Seasonality | Monthly distribution of yearly cases | “Seasonality factor” multiplying expected monthly cases | Monthly distribution of yearly cases |
| VE | Separate VEs for hospitalization and outpatient visit. Waning over 36 months modelled using a cubic polynomial regression model. | Separate VEs for RSV-URTD and RSV-LRTD. First month efficacy reduced by half. Linear waning between months 1-7, 7-18, 18+. | Four VE curves for hospitalization, ED visit, outpatient healthcare provider visit, and non-medically attended RSV. Linear waning between months 0-3, 3-6, 6-12, 12-18, 18-24, 24-36. |
| AEFI | Local and systemic | Local and systemic | Not modelled |
| Vaccination timing | September and October 2024 | October 1, 2024 | September and October 2024 |
| Time horizon | Three years. Third-year VE in two year scenario is assumed to be zero | Any integer | Any integer, but lifetime horizon must be used to capture QALY losses due to RSV mortality |

<sup>a</sup>Not included in results, but cannot be removed from model; <sup>b</sup>RSV upper respiratory tract disease; <sup>c</sup>RSV lower respiratory tract disease Abbreviations: VE = vaccine effectiveness; AEFI=adverse events following immunization

#### PHAC Model

The PHAC model is a static individual-based model with five age groups and two risk strata and includes the following RSV outcomes: outpatient healthcare provider visits, emergency department visits, hospitalization without ICU, hospitalization with ICU, and death (6). Hospitalizations without ICU and hospitalizations with ICU were collapsed to simply hospitalization for comparability with other models. Adverse events following immunization are assumed to result in a proportion of all immunizations given. The model includes lifetime QALY losses for RSV mortality and used a fixed policy time horizon of three years.

#### Pfizer Model

The Pfizer model is a static cohort model with five age groups and two or three risk strata. The model structure has been previously described (14) and the model inputs were adapted for the Canadian context. Because there are fewer age groups in this model than in the policy alternatives considered (described below) we merged some age groups, using population-weighted averages where input assumptions differed between merged age groups.

The model includes costs and QALY losses for non-medically attended cases, outpatient visits, emergency department visits, hospitalizations, and death. We excluded costs and QALY losses associated with non-medically attended cases for consistency with the PHAC model. Adverse events following immunization (AEFIs) are not explicitly considered in this model. We added the expected cost of treating AEFIs ($0.67 per vaccinated person) in the vaccine administration cost, but were unable to incorporate expected QALY losses. Although the model has a user-specified time horizon, a lifetime model horizon is necessary to fully count QALY losses due to RSV mortality. Finally, unlike the other models, the Pfizer model assumes that age-specific parameter inputs are piecewise linear between age groups in one year increments, as illustrated in **Figure 1**.

**Figure 1.**
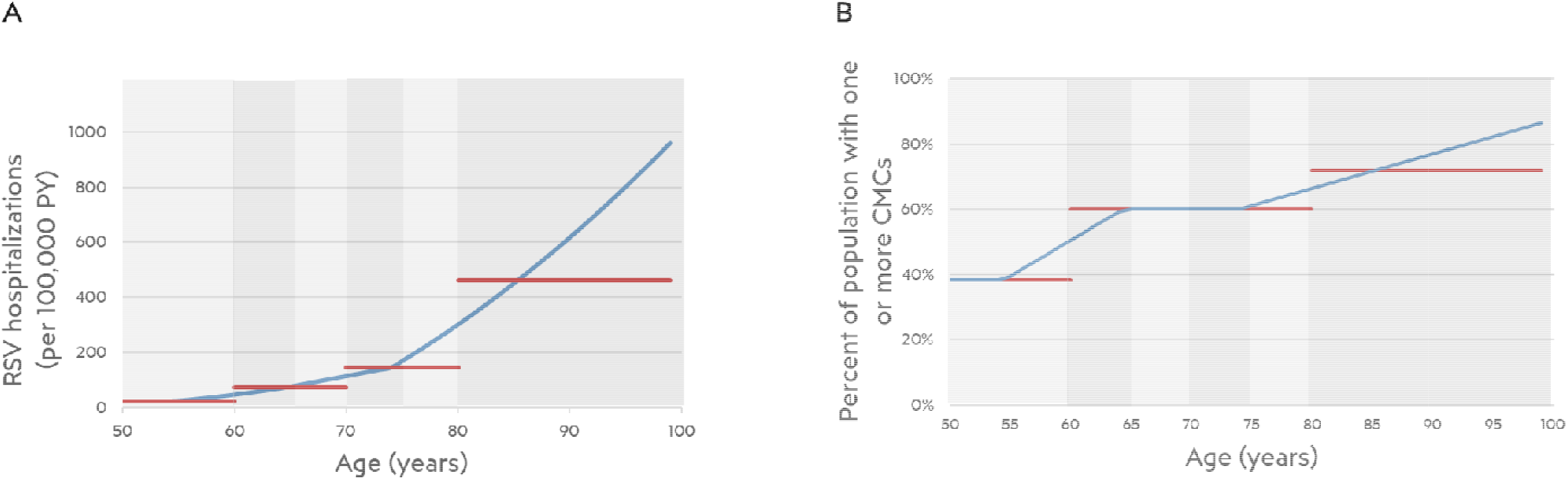
Comparison of Pfizer piecewise linearity (blue) and PHAC/GSK uniform (red) assumptions for age-varying data. Results are shown for (A) incidence of RSV hospitalizations per 100,000 person-years and (B) prevalence of chronic medical conditions.

#### GSK Model

The GSK model is a static cohort model with up to seven age groups. The model structure has been previously described (14, 15) and the model parameters were adapted for the Canadian population. All people in each age group are assumed to be the lowest age; therefore, we staggered age groups to start at the mid-point of desired ranges so that age groups had the same average age and life years lost in the case of RSV mortality. The model does not model risk strata explicitly, but by treating high and low risk people as separate cohorts we achieved the same effect. As in other models, lifetime QALY losses are considered for RSV mortality, but a policy time horizon of two or three years may be selected by the user.

Rather than modeling vaccine effectiveness (VE) as protecting directly against healthcare system use outcomes such as outpatient visits and hospitalization, the original model assumes that all RSV acute respiratory infections (RSV-ARI) lead to either upper respiratory tract disease (RSV-URTD) or lower respiratory tract disease (RSV-LRTD). Differing levels of healthcare resource use are then assumed, depending on whether a person has RSV-URTD or RSV-LRTD. We made a change to this formulation by modeling RSV-LRTD as equivalent to RSV requiring hospitalization, and RSV-URTD as resulting in either outpatient healthcare provider or emergency department visits, with probabilities proportional to the age- and risk-stratified number of cases of each outcome estimated in the PHAC model.

Vaccination in the GSK model confers two levels of protection: against RSV-LRTD and against all RSV-ARI. In the original model VE against RSV-ARI and RSV-LRTD were based on clinical trial results and VE for RSV-URTD was calculated based on the other two VE inputs. For this analysis we computed RSV-ARI waning profiles for each age-risk stratum such that the resulting VE against RSV-URTD matched the assumptions of VE against outpatient and ED visits in the other models.

### Input Parameters

Common input parameters were based on those used in the PHAC cost-utility analysis, which preferentially used Canadian data when available, and otherwise used data from other jurisdictions or expert opinion (6). A full description of input parameters used is published separately (6), and the values used in the current analysis are provided in **Supplementary Table 1** for reference. Some key parameters are described below.

Age-specific proportions of people with one or more CMCs were based on Canadian prevalence estimates of chronic obstructive pulmonary disease, obesity (self-reported body mass index greater than or equal to 30), high blood pressure, cancer, heart disease, suffering from the effects of a stroke, diabetes, or dementia (13). Vaccine coverage was assumed to follow influenza vaccine uptake (16). Vaccination costs included administration costs and the public Canadian list price of $230 per dose for both vaccines. VE against RSV requiring outpatient medical attendance or hospitalization was assumed to be equal to published VE against mild and severe RSV infections, and wane over a two or three year period, with the three-year period estimates based on extrapolation from existing data, which were limited to two RSV seasons at the time of the analysis (17-19). Age-specific incidence of RSV hospitalization was estimated based on results from Canadian studies (2), with an assumed case under-detection factor of 1.5 fold (4). RSV infections were assumed to be seasonal with most cases occurring in January to March (20). Where necessary due to model structural assumptions, we adapted input parameters to have equivalent effects across models, but did not modify the underlying logic of any model.

### Model Comparisons

As described above, we modelled the use of the Abrysvo (Pfizer) or Arexvy (GSK) vaccines separately under the assumption of either two or three years duration of protection following vaccination, with VE assumed to wane over the specified time period. In addition to no vaccination, we evaluated 19 policy alternatives using different combinations of age and comorbidity eligibility requirements under each scenario (6):

- Age-based policies: all adults older than 60, 65, 70, 75, or 80 years of age were considered eligible;
- Medical risk-based policies: all adults older than 60, 65, 70, 75, or 80 years of age who also had one or more chronic medical conditions (CMCs) were considered eligible;
- Age- and medical risk-based policies: all adults over a general age threshold, plus adults with CMCs over a range of lower age thresholds (50 or 60 years) were considered eligible.

While all three models use a lifetime horizon for QALY losses due to RSV mortality, each has differing policy time horizons for evaluating the impact of vaccination programs. As VE is assumed to be finite (i.e., a maximum of three years considered in this analysis), these differing horizons did not impact comparison of incremental cost-effectiveness ratios. In graphical comparisons of cost-effectiveness frontiers, we used net program costs and effects relative to no vaccination to account for differences in model time horizons.

## RESULTS

A comparison of the cost-effectiveness frontiers for the three models using VE assumptions for Abrysvo and Arexvy in the two-year and three-year waning scenarios is shown in **Figure 2**. The three models were broadly concordant across the four scenarios. The PHAC and GSK models identified the same policy alternatives as potentially cost-effective, with the optimal policy dependent on the cost-effectiveness threshold. The Pfizer model had similar results overall, but identified an additional policy, vaccination for higher-risk people over age 65 years, as a potentially cost-effective option. This policy was consistently subject to extended dominance (i.e., not cost-effective at any value of the cost-effectiveness threshold) using the PHAC and GSK models. For all models, all of the policies identified as potentially cost-effective were either risk-based or age- and risk-based; age-based strategies were never identified as cost-effective options. We found that a policy of vaccinating higher-risk people aged 80 years and older dominated a policy of no vaccination across all models using either vaccine’s assumed VE.

**Figure 2.**
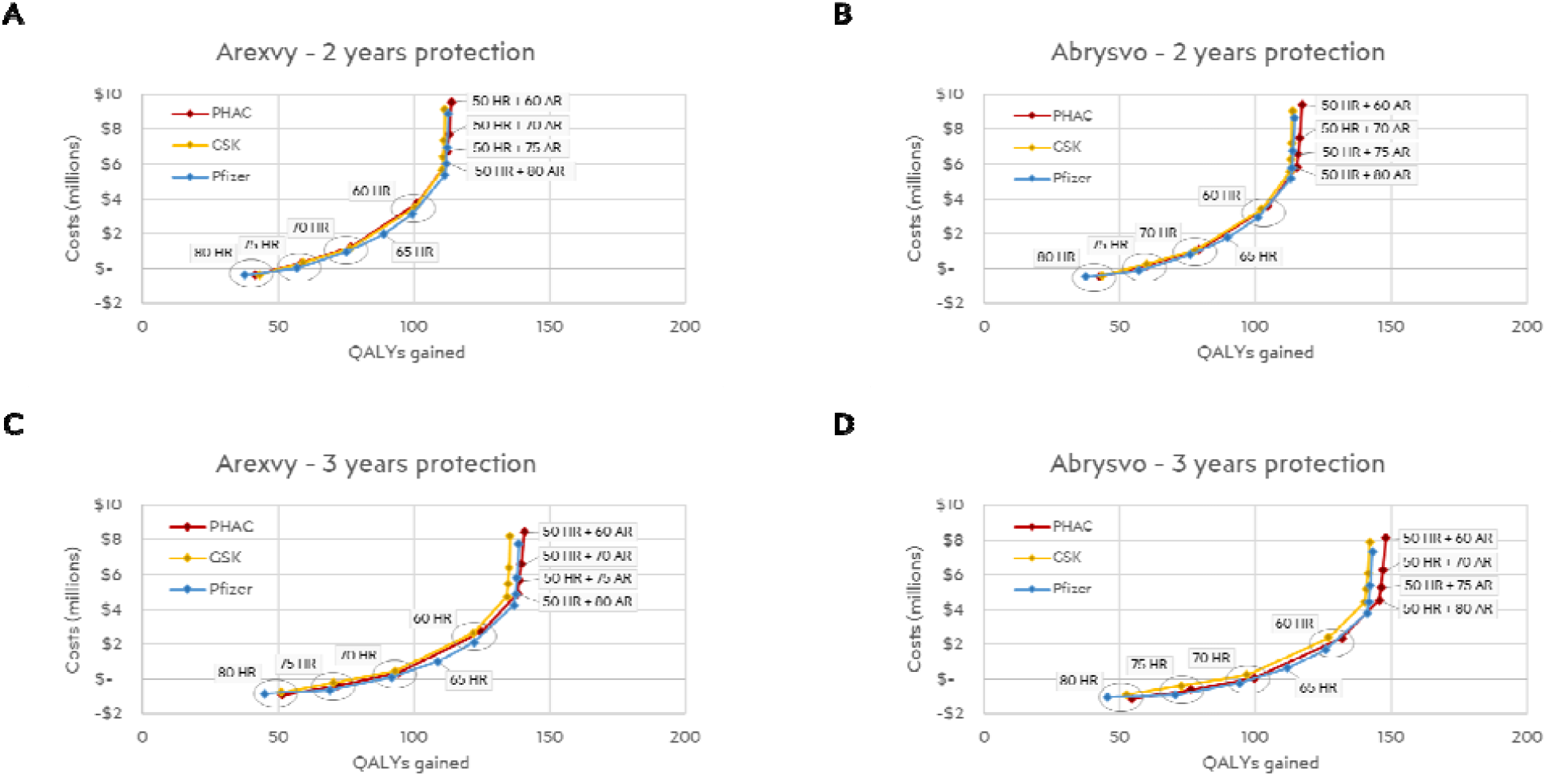
Potentially cost-effective RSV vaccination strategies. Cost-effectiveness frontiers were generated using outputs from the PHAC (red), GSK (yellow), and Pfizer (blue) models. Results are showns for the following scenarios: (A) Arexvy VE data with waning protection over 2 years, (B) Abrysvo VE data with waning over 2 years, (C) Arexvy VE data with waning protection over 3 years, and (D) Abrysvo VE data with waning over 3 years. Labels indicate the vaccination strategy. For clarity, only strategies that were on the cost-effectiveness frontier are shown. All other strategies were dominated or excluded by extended dominance and were not cost-effective options, regardless of the cost-effectiveness threshold used. Incremental cost-effectiveness ratios for the non-dominated strategies are provided in Tables 2 and 3. As described in the methods, costs and QALYs gained are shown relative to no vaccination to allow for a comparison across models. Abbreviations: HR=high risk; AR=average risk; QALY=quality-adjusted life year; VE=vaccine effectiveness.

### Two-year vaccine protection scenarios

Sequential ICERs for all policies that were not dominated or extendedly dominated in the two-year vaccine protection scenario are shown in **Table 2**. Compared to vaccination of higher-risk people aged 80 years and older, the PHAC and GSK models estimated sequential ICERs between $38,029/QALY and $41,325/QALY for a policy of vaccinating higher-risk people over age 75 years, while the Pfizer model had ICERs of approximately $20,000/QALY. With the exception of the Pfizer model parameterized with Arexvy VE estimates, all sequential ICERs for the higher-risk adults age 70 years and older policy were less than the commonly used $50,000/QALY cost-effectiveness threshold when compared to vaccination of higher-risk adults aged 75 years and older; the sequential ICER for the Pfizer model using Arexvy VE estimates was only slightly above this threshold at $50,388/QALY. The Pfizer model was the only model to estimate that a policy of vaccinating higher-risk people over age 65 years could potentially be a cost-effective option, with sequential ICERs of $71,933 to $75,457/QALY compared to a policy for higher-risk adults aged 70 years and older. For all models, sequential ICERs for a policy of vaccinating all higher-risk people over age 60 years were approximately $100,000/QALY compared to vaccination of higher-risk adults aged 70 (PHAC and GSK models) or 65 (Pfizer model) years and older. Above this threshold, the next most cost-effective policies were all those vaccinating higher-risk people over age 50 years and progressively lower age groups of people without CMCs. Strictly age-based policies were never identified as cost-effective options, regardless of the model used.

**Table 2.** Sequential incremental cost-effectiveness ratios ($/QALY) for RSV vaccination strategies identified as potentially cost-effective, assuming that vaccine protection wanes within two years. Results are shown for each model and using data for either the Arexvy or Abrysvo vaccines.

| Policy | Arexvy |  |  | Abrysvo |  |  |
| --- | --- | --- | --- | --- | --- | --- |
|  | PHAC | GSK | Pfizer | PHAC | GSK | Pfizer |
| 80+ HR | - | - | - | - | - | - |
| 75+ HR | \$40,660 | \$41,325 | \$21,219 | \$38,029 | \$39,199 | \$18,682 |
| 70+ HR | \$49,502 | \$48,068 | \$50,388 | \$46,157 | \$45,591 | \$47,309 |
| 65+ HR | Extended dominated | | \$75,457 | Extended dominated | | \$71,933 |
| 60+ HR | \$102,356 | \$99,485 | \$108,641 | \$98,583 | \$96,188 | \$104,544 |
| 80+ AR & 50+ HR | \$214,052 | \$212,578 | \$189,414 | \$209,131 | \$206,540 | \$182,774 |
| 75+ AR & 50+ HR | \$1,317,114 | \$2,865,566 | \$1,329,263 | \$1,421,826 | \$2,784,588 | \$1,265,781 |
| 70+ AR & 50+ HR | \$1,421,897 | \$3,391,567 | \$2,829,476 | \$1,519,503 | \$3,298,674 | \$2,703,549 |
| 60+ AR & 50+ HR | \$2,013,359 | \$5,059,381 | \$3,449,849 | \$2,235,963 | \$4,920,460 | \$3,286,184 |

Only strategies that were not dominated or subject to extended dominance are listed. For this analysis, the ICER is calculated relative to the strategy in the preceding row.
Abbreviations: HR = high risk; AR=average risk

**Table 3.** Sequential incremental cost-effectiveness ratios ($/QALY) for vaccination strategies identified as potentially cost-effective, assuming that vaccine protection wanes within three years. Results are shown for each model and using data for either the Arexvy or Abrysvo vaccines.

| Policy | Arexvy |  |  | Abrysvo |  |  |
| --- | --- | --- | --- | --- | --- | --- |
|  | PHAC | GSK | Pfizer | PHAC | GSK | Pfizer |
| 80+ HR | - | - | - | - | - | - |
| 75+ HR | \$26,834 | \$27,801 | \$8,269 | \$23,169 | \$24,695 | \$4,745 |
| 70+ HR | \$32,907 | \$29,320 | \$32,736 | \$28,814 | \$25,727 | \$28,557 |
| 65+ HR | Extended dominated | | \$53,647 | Extended dominated | | \$48,856 |
| 60+ HR | \$77,338 | \$76,430 | \$81,335 | \$71,776 | \$71,513 | \$75,674 |
| 80+ AR & 50+ HR | \$165,771 | \$169,258 | \$147,249 | \$158,601 | \$153,336 | \$138,356 |
| 75+ AR & 50+ HR | \$1,090,178 | \$2,350,773 | \$975,340 | \$1,135,141 | \$1,461,416 | \$905,774 |
| 70+ AR & 50+ HR | \$1,219,926 | \$2,534,807 | \$2,132,683 | \$1,290,424 | \$1,560,714 | \$1,984,098 |
| 60 AR+ & 50+ HR | \$1,588,568 | \$4,112,609 | \$2,577,733 | \$1,717,449 | \$2,547,744 | \$2,394,941 |

Only strategies that were not dominated or subject to extended dominance are listed. Note that for the results in this table, the ICER is calculated relative to the strategy in the preceding row.
Abbreviations: HR = high risk; AR=average risk

### Three-year vaccine protection scenarios

Sequential ICERs for all policies that were not dominated or extendedly dominated for a scenario assuming that vaccine protection extends through a third season are provided in **Table 3**. ICERs for these scenarios were predictably lower than their equivalents in the two-year scenarios, owing to longer assumed duration of vaccine protection. Sequential ICERs for a policy of vaccinating higher-risk people over age 70 years were between $25,727/QALY and $32,907/QALY compared to vaccination of higher-risk adults over age 80 years. As with the two-year vaccine protection scenario, only the Pfizer model identified vaccinating higher-risk people over age 65 years as a cost-effective option, with sequential ICERs of $48,856 to $53,647/QALY compared to a policy for higher-risk adults aged 70 years and older. Vaccinating higher-risk people over age 60 years resulted in ICERs between $71,513/QALY and $81,335/QALY compared to vaccination for higher-risk adults aged 70 (PHAC and GSK models) or 65 (Pfizer) years and older. At higher cost-effectiveness thresholds, as with the two-year vaccine protection scenarios, age- and risk-based policies that included vaccinating higher-risk people over age 50 years and progressively lower age groups of average risk people were identified as cost-effective options. Age-based policies were never cost-effective when compared with these other policy options.

### Age-based policies

Though age-based policies were never identified as cost-effective when considered alongside risk- or age- and risk-based options, these may be preferred by some decision-makers based on other considerations, such as potentially reduced complexity of program delivery. We therefore performed a sub-analysis of the two-year vaccine protection scenarios, restricted to only age-based policies (**Table 4**). Sequential ICERs for a policy of vaccinating all people over age 80 years were between $3,161/QALY and $6,194/QALY compared to no vaccination. Policies including younger people were unlikely to be considered cost-effective at a $50,000/QALY cost-effectiveness threshold; the PHAC and GSK models estimated ICERs between $78,637/QALY and $85,805/QALY for a policy of vaccinating all people over age 75 years compared to a policy for all people over age 80 years. However, the Pfizer model had ICERs of between $50,090/QALY and $53,205/QALY in this scenario. More expansive age-based policies had progressively higher ICERs and were unlikely to be considered cost-effective at commonly-used thresholds.

**Table 4.** Sequential incremental cost-effectiveness ratios ($/QALY) comparing only age-based strategies and assuming vaccine protection wanes within two years. Results are shown for each model and using data for either the Arexvy or Abrysvo vaccines.

| Policy | Arexvy |  |  | Abrysvo |  |  |
| --- | --- | --- | --- | --- | --- | --- |
|  | PHAC | GSK | Pfizer | PHAC | GSK | Pfizer |
| No vaccination | -- | -- | -- | -- | -- | -- |
| 80+ All | \$5,391 | \$6,194 | \$5,883 | \$3,261 | \$4,838 | \$3,161 |
| 75+ All | \$82,326 | \$85,805 | \$53,205 | \$78,637 | \$82,607 | \$50,090 |
| 70+ All | \$99,045 | \$100,332 | \$100,829 | \$94,264 | \$96,651 | \$96,496 |
| 65+ All | Extended dominated | | \$136,241 | Extended dominated | | \$131,165 |
| 60+ All | \$172,061 | \$172,531 | \$201,336 | \$167,226 | \$167,515 | \$194,749 |

Note that for the results in this table, the ICER is calculated relative to the strategy in the preceding row.

## DISCUSSION

Our multi-model comparison of three Canadian cost-utility models has shown that RSV vaccination programs for older adults may be a cost-effective intervention, particularly when these programs are focused on population groups with the highest risk of RSV disease. These findings were broadly concordant across the scenarios considered; the policies identified as optimal at commonly-used cost-effectiveness thresholds were generally consistent. Additionally, estimated ICERs did not differ greatly between the two vaccines considered.

Using harmonized model input parameters, all models consistently identified policies based on medical risk as optimal compared to policies based only on age. One difference across the models related to the identification of a policy of vaccinating higher-risk adults over age 65 years as potentially cost-effective only using the Pfizer model. Using the other two models, this policy option was extendedly dominated, as there were alternative policy options that provided better value for money. This difference is likely due how the models use age-varying data. Although the assumption in the PHAC and GSK models of constant values across each age grouping aligns more closely with source data, the age gradient assumptions used by the Pfizer model may be considered more realistic by some decision-makers.

Although most other published economic evaluations of RSV vaccination in older adults to date have focused on age-based strategies only, the general trends observed in our analysis can be compared with other studies. A systematic review of economic evaluations of RSV vaccines in adults conducted in the United States and Hong Kong found that in most studies vaccination programs offered to all adults aged 60 or 65 years and older were unlikely to be cost-effective using a $50,000/QALY threshold, unless there was a substantial reduction in vaccine price (21). As with our analysis, studies that considered multiple age cutoffs for vaccination programs found that ICERs were lower when programs were more restrictive with respect to age eligibility (14, 22). A recent Canadian economic evaluation examined policies offering vaccine to residents of long-term care homes alone or alongside age-based vaccination of community dwelling adults (23). This study used a threshold analysis to identify the maximum vaccine price at which vaccination would be cost-effective for a $50,000/QALY threshold and found that higher vaccine prices were acceptable for vaccination strategies restricted to residents of long-term care homes, where the risk of RSV disease is highest. The maximum acceptable vaccine price was reduced as age eligibility for community-dwelling adults was expanded to younger ages (23).

This analysis has some limitations which must be considered when interpreting our results. All models included in our comparison were static models and did not consider indirect effects of vaccination programs. As a result, these models may underestimate the potential cost and QALY savings of these programs, leading to the identification of less expansive policy options as optimal. Second, we did not conduct an analysis using the societal perspective and did not consider the possible impact of vaccination for the prevention of non-medically-attended RSV disease; our findings may underestimate the benefits of vaccination programs. Finally, we limited our analysis to a small number of scenarios and did not conduct sensitivity analyses. However, given the consistency of our results across the models, the value of further exploration of the impact of parameter uncertainty is likely small for this comparative analysis.

In conclusion, our multi-model comparison shows that RSV vaccination programs are likely cost-effective for some subgroups of older Canadian adults, particularly those with CMCs that place them at increased risk of RSV disease. These findings are robust to alternate model structural assumptions.

## Data Availability

All data produced in the present study are available upon reasonable request to the authors.

## Acknowledgements

The GSK Canadian cost-utility model for older adults was developed by Sydney George (GSK, Mississauga, Canada), Michael Dolph (Cytel, Inc., Toronto, Canada), Yufan Ho (GSK, Singapore), Dessi Loukov (GSK, Mississauga, Canada), Emily Matthews (Cytel, Inc., Toronto, Canada), Shreena Malaviya (Cytel, Inc., Toronto, Canada), Janine Xu (GSK, Mississauga, Canada), Daniel Molnar (GSK, Wavre, Belgium). The Pfizer Canadian cost-utility model for adults was developed by Ahuva Averin (Avalere Health, Boston, USA), Mark Atwood (Avalere Health, Boston, USA), Derek Weycker (Avalere Health, Boston, USA), Erin Quinn (Avalere Health, Boston, USA), Alexandra Goyette (Pfizer Canada Inc., Kirkland, Canada), Reiko Sato (Pfizer Inc., New York, USA). The authors thank members of National Advisory Committee on Immunization RSV Working Group, who provided feedback on model parameters.

