## Supplementary Table 1 for "Cost-effectiveness of respiratory syncytial virus vaccination strategies for older Canadian adults: a multi-model comparison"

**Supplementary Table 1**. Model input parameters from Tuite et al. 2024 (1) used in the present analysis.

| Parameter | Base | Range | Reference |
| --- | --- | --- | --- |
| Population distribution (%) | | | |
| 50-59 years | 31.7 | -- | Statistics Canada (2) |
| 60-64 years | 17.5 | -- |  |
| 65-69 years | 15.8 | -- |  |
| 70-74 years | 12.8 | -- |  |
| 75-79 years | 9.9 | -- |  |
| 80+ years | 12.2 | -- |  |
| % of population with one or more CMCs | | | |
| 50-59 years | 38.4 | -- | Statistics Canada (3) |
| 60-79 years | 60.1 | -- |  |
| 80+ years | 72.1 | -- |  |
| Monthly % of annual RSV cases | | | |
| September | 1.2 | -- | Respiratory Virus Detection Surveillance System (average of 9 seasons, 2010-2011 to 2018-2019) (4) |
| October | 1.9 | -- |  |
| November | 5.5 | -- |  |
| December | 14.2 | -- |  |
| January | 17.6 | -- |  |
| February | 21.1 | -- |  |
| March | 17.0 | -- |  |
| April | 11.0 | -- |  |
| May | 5.6 | -- |  |
| June | 2.6 | -- |  |
| July | 1.3 | -- |  |
| August | 1.1 | -- |  |
| Odds ratio for medically-attended outpatient care in adults with one or more CMCs | | | |
| All ages | 1.1 | -- | Shi et al. 2022 (5) |
| % of patients requiring hospitalization with one or more CMCs | | | |
| All ages | 98.2 | -- | ElSherif et al. 2023 (6) |
| Under-detection factor for medically-attended RSV in adults | | | |
| All ages | 1.5 | 1-2 | McLaughlin et al. 2022 (7) |
| Annual incidence of medically-attended RSV requiring outpatient healthcare provider visit per 100,000 population (unadjusted for under-detection) | | | |
| 50-59 years | 261.9 | 186.5-337.3 | ElSherif et al. 2023 (6); McLaughlin et al. 2022 (7); Respiratory Virus Detection Surveillance System (4) |
| 60-69 years | 604.1 | 472.8-707.8 |  |
| 70-79 years | 780.0 | 625.1-934.1 |  |
| 80+ years | 2487.1 | 2097.1-2877.2 |  |
| Annual incidence of medically-attended RSV requiring emergency department visit per 100,000 population (unadjusted for under-detection) | | | |
| 50-59 years | 16.8 | 12.0-21.6 | ElSherif et al. 2023 (6); McLaughlin et al. 2022 (7); Respiratory Virus Detection Surveillance System (4) |
| 60-69 years | 43.3 | 33.9-50.7 |  |
| 70-79 years | 68.5 | 54.9-82.0 |  |
| 80+ years | 218.3 | 184.1-252.5 |  |
| Annual incidence of RSV-attributable hospitalization per 100,000 population (unadjusted for under-detection) | | | |
| 50-59 years | 15.1 | 10.8-19.5 | ElSherif et al. 2023 (6); Respiratory Virus Detection Surveillance System (4) |
| 60-69 years | 47.5 | 37.2-55.7 |  |
| 70-79 years | 96.4 | 77.3-115.4 |  |
| 80+ years | 307.4 | 259.2-355.6 |  |
| % of patients hospitalized with RSV requiring ICU admission | | | |
| All ages | 13.7 | 10.2-17.9 | ElSherif et al. 2023 (6) |
| % of patients with medically attended RSV prescribed an antimicrobial | | | |
| All ages | 50 | 14-89 | Bernardo et al. 2019 (8); ElSherif et al. 2023 (6) |
| RSV mortality per hospitalization (%) | | | |
| 50-64 years | 7.2 | 5.4-9.5 | Chen et al. 2024 (9) |
| 65-74 years | 6.6 | 5.2-8.4 |  |
| 75+ years | 10.1 | 9.0-11.3 |  |
| All-cause mortality rate (per year, per 1,000 population) | | | |
| All ages | Age-specific rates | -- | Statistics Canada (10) |
| Immunization coverage (%), with chronic medical conditions | | | |
| 50-59 years | 58.6 | -- | Seasonal Influenza Vaccination Coverage Survey, 2022-2023 (11) |
| 60-64 years | 59.9 | -- |  |
| 65-69 years | 65.2 | -- |  |
| 70-79 years | 82.7 | -- |  |
| 80+ years | 83.4 | -- |  |
| Immunization coverage (%), without chronic medical conditions | | | |
| 50-59 years | 36.7 | -- | Seasonal Influenza Vaccination Coverage Survey 2022-2023 (11) |
| 60-64 years | 49.4 | -- |  |
| 65-69 years | 61.1 | -- |  |
| 70-79 years | 74.9 | -- |  |
| 80+ years | 74.8 | -- |  |
| Vaccine effectiveness (%), Arexvy (GSK) | | | |
| Outpatient RSV – season 1  (7 mo follow-up) | 82.6 | -- | Friedland 2023 (12); Ison et al. 2024 (13); assumption for season 3 |
| Outpatient RSV – season 2  (6 mo follow-up) | 56.1 | -- |  |
| Outpatient RSV – season 3 | 18.7 | -- |  |
| Hospitalized RSV – season 1  (7 mo follow-up) | 94.1 | -- |  |
| Hospitalized RSV – season 2  (6 mo follow-up) | 64.2 | -- |  |
| Hospitalized RSV – season 3 | 21.4 | -- |  |
| Vaccine effectiveness (%), Abrysvo (Pfizer) | | | |
| Outpatient RSV – season 1  (7 mo follow-up) | 65.1 | -- | Gurtman 2023 (14); assumption for season 3 |
| Outpatient RSV – season 2  (4 mo follow-up) | 48.9 | -- |  |
| Outpatient RSV – season 3 | 16.3 | -- |  |
| Hospitalized RSV – season 1  (7 mo follow-up) | 88.9 | -- |  |
| Hospitalized RSV – season 2  (4 mo follow-up) | 78.6 | -- |  |
| Hospitalized RSV – season 3 | 26.2 | -- |  |
| Vaccine wastage rate (%) | | | |
| All ages | 5 | -- | WHO, 2019 (15) |
| Adverse events following immunization (%) | | | |
| Severe local adverse event | 0.51 | 0.16-1.84 | Melgar et al. 2023 (16) |
| Severe systemic adverse event | 0.57 | 0.10-2.35 |  |
| Cost of vaccine administration per dose ($) | | | |
| All ages | 18 | 13-22 | O’Reilly et al. 2017 (17) |
| Immunization cost per dose ($) | | | |
| GSK | 230 | 100-230 | Robertson, 2023 (18) |
| Pfizer | 230 | 100-230 |  |
| Attributable costs per person hospitalized with RSV ($) | | | |
| Hospitalization (6 months) | 32,228 | 31,622-32,836 | Mac et al. 2023 (19) |
| Hospitalization, die in hospital | 27,534 | 22,027-33,041* |  |
| Costs per person with RSV treated in the outpatient setting ($) | | | |
| Healthcare provider visit | 62 | 48-82 | Sander et al. 2010 (20); CIHI (21); Alliance for Healthier Communities (22) |
| ED visit | 340 | 302-509 |  |
| Direct medical costs for severe local adverse event following vaccination ($) | | | |
| <65 years | 62 | 48-82 | Sander et al. 2010 (20); CIHI (21); Lee et al. 2009 (23) |
| 65+ years | 63 | 49-83 |  |
| Direct medical costs for severe systemic adverse event following vaccination ($) | | | |
| <65 years | 62 | 48-82 | Sander et al. 2010 (20); CIHI (21); Lee et al. 2009 (23) |
| 65+ years | 66 | 51-87 |  |
| Transportation costs ($) | | | |
| Cost of travel to inpatient care | 417 | 210-623 | NACI (24) |
| Background health utility | | | |
| 50-59 years | 0.848 | -- | Yan et al. 2023 (25) |
| 60-64 years | 0.839 | -- |  |
| 65-74 years | 0.867 | -- |  |
| 75+ years | 0.861 | -- |  |
| QALY loss, outpatient, with or without ED visit | | | |
| All ages | 0.0056 | 0.0037 – 0.0075 | Herring et al. 2022 (26); Mao et al. 2022 (27); Zeevat et al. 2022 (28); Meijboom et al. 2013 (29) |
| QALY loss, hospitalization | | | |
| All ages | 0.020 | 0.017-0.030 | Herring et al. 2022 (26); Mao et al. 2022 (27); Zeevat et al. 2022 (28); Meijboom et al. 2013 (29) |
| QALY loss, death | | | |
| 50-59 years | 20.26 | -- | Yan et al. 2023 (25); Statistics Canada (10, 30) |
| 60-64 years | 16.74 | -- |  |
| 65-69 years | 14.29 | -- |  |
| 70-74 years | 11.75 | -- |  |
| 75-79 years | 9.38 | -- |  |
| 80+ years | 5.84 | -- |  |
| QALY loss, adverse event following vaccination | | | |
| Serious local adverse event | 0.0003 | 0.0002-0.0004 | Prosser et al. 2023 (31); assumption |
| Serious systemic adverse event | 0.0004 | 0.0003-0.0005 |  |

*Range defined as ±20% of the base value
